# Sex-specific associations of adiposity with cardiometabolic traits: multi-life-stage cohort study with repeat metabolomics

**DOI:** 10.1101/2020.11.30.20240895

**Authors:** Linda M. O’Keeffe, Joshua A. Bell, Kate N. O’Neill, Matthew Lee, Mark Woodward, Sanne Peters, George Davey Smith, Patricia M. Kearney

## Abstract

**Background:** Sex differences in cardiometabolic disease risk are commonly observed across the life course but are poorly understood and may be due to different cardiometabolic consequences of adiposity in females and males. We examined whether adiposity influences cardiometabolic trait levels differently in females and males at four different life stages.

**Methods:** Data were from two generations (offspring, Generation 1 [G1] and their parents, Generation 0 [G0]) of the Avon Longitudinal Study of Parents and Children birth cohort study. Body mass index (BMI) and total fat mass from dual-energy x-ray absorptiometry were measured at mean age 9y, 15y and 18y in G1. Waist circumference was measured at 9y and 15y in G1. Concentrations of 148 cardiometabolic traits quantified using nuclear magnetic resonance spectroscopy were measured at 15y, 18y and 25y in G1. In G0, all three adiposity measures and the same 148 traits were available at 50y.Using linear regression models, sex-specific associations of adiposity measures at each time point (9y, 15y and 18y) with cardiometabolic traits 3 to 6 years later were examined in G1. In G0, sex-specific associations of adiposity measures and cardiometabolic traits were examined cross-sectionally at 50y.

**Results:** 3081 G1 and 4887 G0 participants contributed to analyses. BMI was more strongly associated with key atherogenic traits in males at younger ages (15y-25y) and associations were more similar between the sexes or stronger in females at 50y, particularly for apolipoprotein-B-containing lipoprotein particles and lipid concentrations. For example, a 1- SD (3.8 kg/m^2^) higher BMI at 18y was associated with 0.36 SD (95% Confidence Interval (CI) = 0.20, 0.52) higher concentrations of extremely large very-low-density lipoprotein (VLDL) particles at 25y in males compared with 0.15 SD (95% CI = 0.09, 0.21) in females. In contrast, at 50y, a 1-SD (4.8 kg/m^2^) higher BMI was associated with 0.33 SD (95% CI = 0.25, 0.42) and 0.30 SD (95% CI = 0.26, 0.33) higher concentrations of extremely large VLDL particles in males and females respectively. Sex-specific associations of DXA-measured fat mass and waist circumference were similar to findings for BMI in both generations and at all ages.

**Conclusion:** The results of this study suggest that the adverse cardiometabolic effects of adiposity are stronger and begin earlier in the life course among males compared with females until mid life, particularly for key atherogenic lipids. Adolescent and young adult males may therefore be high priority targets for obesity prevention efforts.

## Introduction

Cardiometabolic diseases including type 2 diabetes mellitus (T2DM) and cardiovascular disease (CVD) are a leading cause of death worldwide (1). Females and males do not experience this risk equally; for instance, males develop higher systolic blood pressure (SBP) during adolescence and this sex difference widens in early adulthood and persists throughout much of the life course (2-4). Furthermore, age-adjusted CVD risk is higher in males compared with females throughout adulthood, and although this sex difference attenuates with age, female and male CVD risk only become similar from approximately the 7^th^ or 8^th^ decade onwards (5). The underlying aetiology of this sex difference remains poorly understood, however, and opportunities to develop and deliver more effective sex-specific CVD prevention across the life course are under-explored.

Emerging evidence suggests that the cardiometabolic consequences of adiposity may differ in females and males (6, 7), contributing to sex differences in cardiometabolic disease risk. To date, several studies have examined sex-specific associations of adiposity with clinical endpoints such as coronary heart disease (CHD) in adults (8-16), although results have been conflicting and based mostly on BMI. In contrast, few studies have examined sex-specific associations of adiposity with cardiometabolic trait measures from metabolomics platforms that provide more granular insight into CVD aetiology (7). Moreover, the cardiometabolic consequences of adiposity among females and males across different life stages has not been investigated (8-16), despite potentially important implications for the timing of disease prevention across the life course.

Using data from two generations of the Avon Longitudinal Study of Parents and Children (ALSPAC) birth cohort, we compared the sex-specific associations of body mass index (BMI), waist circumference and dual-energy x-ray absorptiometry (DXA)-determined total fat mass with cardiometabolic traits from targeted metabolomics measured three times in offspring (Generation 1 cohort [G1] in mid adolescence, late adolescence and early adulthood) and once in mid life in their parents (Generation 0 cohort [G0]).

## Methods

### Study population

Data were from participants of ALSPAC, a population-based birth cohort study in which 14,541 pregnant women expected to deliver between 1 April 1991 and 31 December 1992 were recruited from the former county of Avon in southwest England. Offspring (G1 cohort) alive at 1 year (n=13,988) have been followed up with multiple assessments with an additional 913 children enrolled over the course of the study (17, 18). Parents (G0 cohort) of G1 offspring participants have also been followed with multiple assessments (19). Parental data used here were primarily from mothers who attended a clinic assessment between December 2008 and July 2011 and from fathers who attended a clinic assessment between September 2011 and February 2013. Study data among G1 cohort participants after age 22 years were collected and managed using REDCap electronic data capture tools hosted at University of Bristol (20, 21).

Ethical approval was obtained from the ALSPAC Law and Ethics and Local Research Ethics Committees. The study website contains details of all the data that is available through a fully searchable data dictionary and variable search tool (http://www.bristol.ac.uk/alspac/researchers/our-data/).

## Data

### Assessment of adiposity

In the G1 cohort, BMI and DXA-determined total fat mass were measured at mean age 9y, 15y and 18y, with the additional measurement of waist circumference at 9y and 15y. In the G0 cohort, all three measures were available at 50y. Weight was recorded to the nearest 0.1 kg using a Tanita scale. Height was measured in light clothing without shoes to the nearest 0.1 cm using a Harpenden stadiometer. BMI was calculated as weight (in kilograms) divided by the square of height (in meters). Waist circumference was measured using a flexible tape to measure circumference to the nearest 1 mm at the mid-point between the lower ribs and the pelvic bone. Total body fat mass (in kg, less head) was derived from whole body DXA scans using a GE Lunar Prodigy (Madison, WI, USA) narrow fan beam densitometer.

### Assessment of cardiometabolic traits

In the G1 cohort, blood samples were drawn in clinics at ages 15y, 18y and 25y. Bloods were taken after a minimum of a 6-hour fast. Proton nuclear magnetic resonance (^1^H-NMR) spectroscopy from a targeted metabolomics platform (22) was performed on EDTA-plasma samples from each of these three occasions to quantify 148 metabolite concentrations including cholesterol, triglyceride, and other lipid content in lipoprotein subclass particles, apolipoproteins, fatty acids, amino acids, and inflammatory glycoprotein acetyls.

In the G0 cohort, blood samples were drawn in clinics at mean 48y in mothers and mean 53y in fathers (combined mean (SD) age of 50y (5y)). Bloods were taken after a minimum of a 6-hour fast. The ^1^H-NMR metabolomics platform used in the G1 cohort was performed on serum samples taken on the G0 cohort to quantify the same 148 metabolite concentrations. In mothers only, additional blood samples were available from visits when mothers were aged 51y, from which additional measures of the same 148 cardiometabolic concentrations were quantified. In this study, if mothers had missing data on all traits on the first measurement occasion (age 48y) but had data on at least one trait on the second measurement occasion (51y), the trait from the second occasion was used to replace missing values on the first occasion. The number of mothers with these replacements ranged from 215 to 223 across traits.

### Assessment of confounders

In the G1 cohort analyses, we adjusted for age at clinic completion, ethnicity, education of the child’s mother and father, maternal smoking during pregnancy, birthweight, gestational age, maternal age, parental household social class measured close to the time of offspring birth and height and height^2^ at the time of exposure measurement. In analyses which included trait concentrations at 18y and 25y as outcomes, we also additionally adjusted for G1 offspring smoking. In the G0 cohort analyses, we adjusted for age at clinic completion, ethnicity, own education, smoking during G1 cohort pregnancy, own social class measured during the G0 child’s pregnancy and height and height^2^ at the time of exposure measurement. Further details on the measurement of confounders are included in eAppendix 1 of Supplementary Material.

### Participants eligible for analysis

Adiposity measures at 9y, 15y and 18y were examined in relation to cardiometabolic traits one occasion after (∼3 to 6 years later at 15y, 18y and 25y) in G1. Adiposity measures at 50y in G0 were examined in relation to cardiometabolic traits also measured at 50y. To allow full use of measured data, analyses were conducted using maximum numbers of participants (with N varying across ages and between traits). Participants were eligible for inclusion in analyses at any age if they had data on all adiposity measures, sex, age, and at least one of the cardiometabolic traits plus all relevant potential confounders as listed above. In the G1 cohort, this resulted in 2190 eligible offspring (1121 females, 1069 males) contributing to analysis of adiposity at 9y and traits at 15y, 1600 (813 females, 787 males) contributing to analyses of adiposity at 15y and traits at 18y and 1613 contributing (928 females, 685 males) to analyses of adiposity at 18y and traits at 25y. The single time-point analysis in G0 parents included 4887 eligible parents (3446 mothers, 1441 fathers). A flow diagram of the study design is shown in Figure 1.

**Figure 1.**
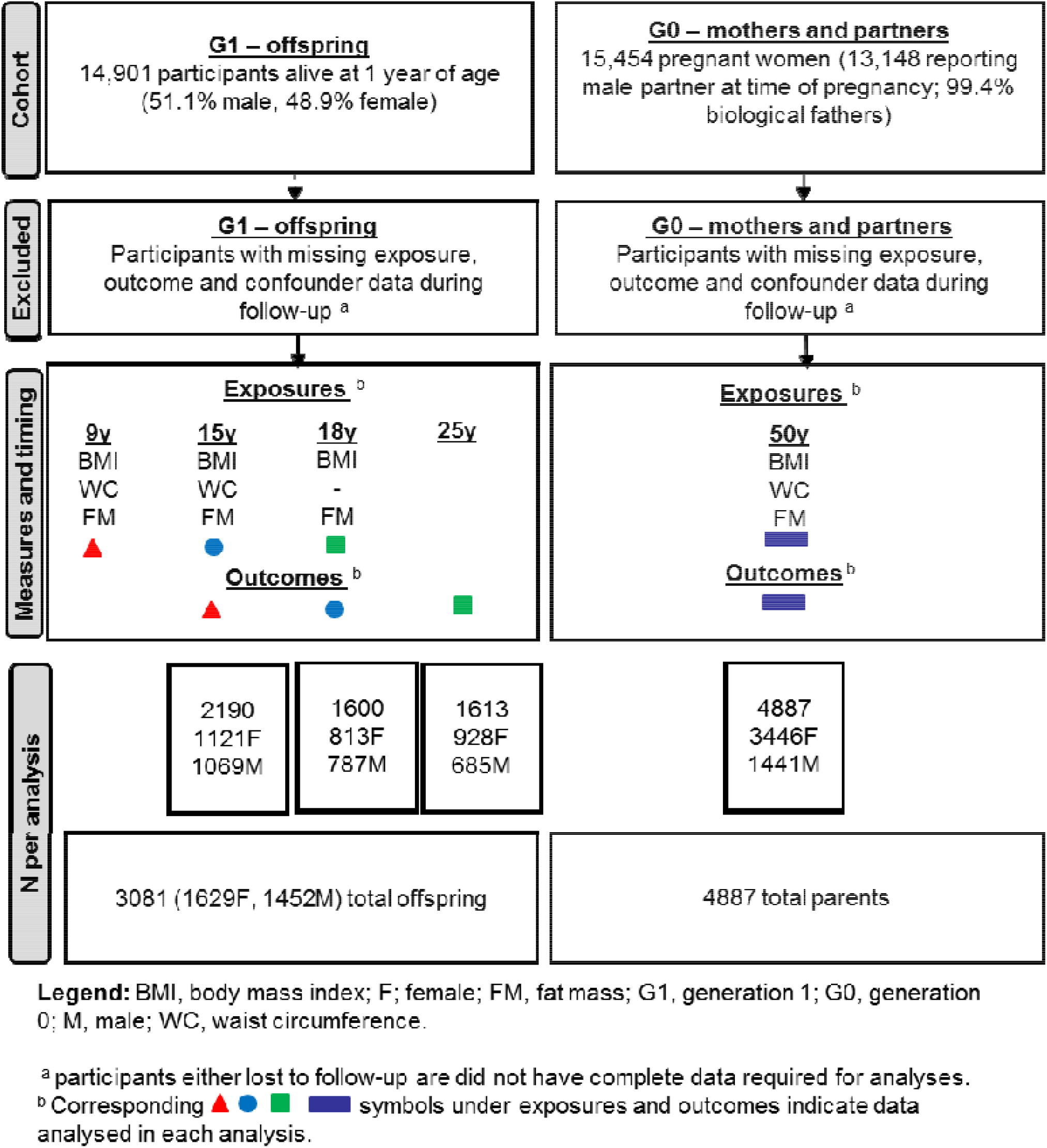
Flow diagram of study design.

### Statistical analysis

#### Primary analysis

We standardised all measures of adiposity and all cardiometabolic traits into standard deviation (SD) units by generating z scores (subtracting the sex-combined mean and dividing by the SD). Linearity of associations was also examined by comparing models where fifths of adiposity measures were treated as continuous exposures to models where fifths of adiposity measures were treated as categorical exposures using a likelihood ratio (LR) test. Mean traits by fifths of exposure were also plotted and examined to explore substantial deviations from linearity. We found no consistent evidence of departures from linearity for most traits across time points (results from LR tests for key traits shown in eTable1) and analyses here therefore treat adiposity measures as continuous exposures. Correlations between adiposity measures were also examined and all three measures were highly correlated with each other in each sex at each age with Pearson correlations of greater than 0.8/0.9 between measures.

Linear regression models with robust standard errors to accommodate skewed outcome distributions were used to examine the association between each standardized adiposity measure (BMI, waist circumference and fat mass) and each standardized cardiometabolic trait. These models included an interaction term for sex and the standardised adiposity measure; this provided the sex-specific mean difference and 95% confidence interval (CI) in each standardized cardiometabolic trait per standard deviation increase in the adiposity measure. All analyses were performed as described above (inclusive of sex-interactions), initially unadjusted, with subsequent adjustment for potential confounders. We also fitted a sex interaction term with all confounding variables separately to allow confounding structures to differ in females and males. As recommended by the American Statistical Association and others (23-25), we present exact P-values (in Supplementary Material) and focus our descriptions of results on effect size and precision.

Analyses were conducted using Stata 15.1 (StataCorp, College Station, Texas, USA) and data visualisation was performed in R (version 3.6.3) using the ggforestplot (0.0.2) package.

#### Sensitivity and additional analyses

We repeated all analyses using cardiometabolic traits in their original (non-SD) units to aid clinical interpretation. We repeated our analysis using adiposity measures that were standardised using the sex-specific mean and SD to examine whether results were appreciably different to our main analytic approach (which used sex-combined means and SDs for standardisation). We repeated all analyses on the sample whose full family units were eligible for inclusion in all analyses, i.e. offspring had to be eligible for inclusion in each of the three G1 analyses and mothers and fathers had to be eligible for inclusion in the G0 analysis. We also repeated our analyses excluding participants in the top fifth of the adiposity distribution at each time point to exclude the possibility of results reflecting threshold effects of adiposity measures on cardiometabolic risk. In order to examine potential selection bias in our analysis of G0, we compared characteristics of the G1 participants included in our analyses (n=3081) compared with G1 participants excluded from analyses by sex. Similarly, we examined these characteristics in G0 participants included in our analyses (n=4887) compared with G0 participants excluded from analyses by sex. For both G1 and G0 analyses comparing included and excluded participants, we used maternal and paternal socio-demographic and health characteristics measured at or close to birth (at the commencement of the cohort in 1991/1992 when missing data for most characteristics was rarest).

## Results

### Characteristics of included vs. excluded participants

G1 offspring included in the analysis had a smaller proportion of non-white ethnicity, had parents with higher educational attainment, higher household social class, and mothers with lower levels of smoking during pregnancy compared with offspring excluded from analyses (eTable 2). G1 offspring included had older mothers during pregnancy compared with offspring excluded from analyses, though included and included participants were similar in birthweight, gestational age and adiposity at each age.

G0 parents included in analyses had a smaller proportion of non-white ethnicity, were more educated, had higher social class, lower levels of smoking during pregnancy and were less adipose than parents excluded from analyses (eTable 3).

### Characteristics of participants included in analyses by sex and generation

A minority of male G1 participants and female G1 participants (each < 5.0%) were of non-white ethnicity. Male G1 participants had parents with slightly higher education, lower levels of smoking among their mothers during pregnancy, higher household social class and higher birthweight but similar gestational age and maternal age compared with female G1 participants. Male G1 participants had similar BMI and waist circumference but lower fat mass at each age compared with female participants.

A low proportion of G0 fathers and mothers (each < 5.0%) were of a non-white ethnicity. G0 fathers had higher educational levels and social class and higher levels of smoking during their partners’ pregnancy compared with G0 mothers. G0 fathers had higher BMI and waist circumference but lower fat mass compared with G0 mothers.

### Associations of adiposity measures with lipoprotein concentrations in females and males

In confounder adjusted analyses, adiposity measures (BMI, waist circumference and fat mass) were positively associated with all concentrations of extremely large VLDL particles in both sexes (Figure 2 and eTable 5); associations at 15y, 18y and 25y were stronger in males but more similar between the sexes at 50y. For instance, a 1-SD (3.8 kg/m^2^) higher BMI at 18y was associated with 0.36 SD (95% Confidence Interval (CI) = 0.20, 0.52) higher concentrations of extremely large VLDL particles at 25y in males compared with 0.15 SD (95% CI = 0.09, 0.21) in females. In contrast, at age 50y, a 1-SD (4.8 kg/m^2^) higher BMI was associated with 0.33 SD (95% CI = 0.25, 0.42) and 0.30 SD (95% CI = 0.26, 0.33) higher concentrations of extremely large VLDL particles in males and females respectively.

**Figure 2:**
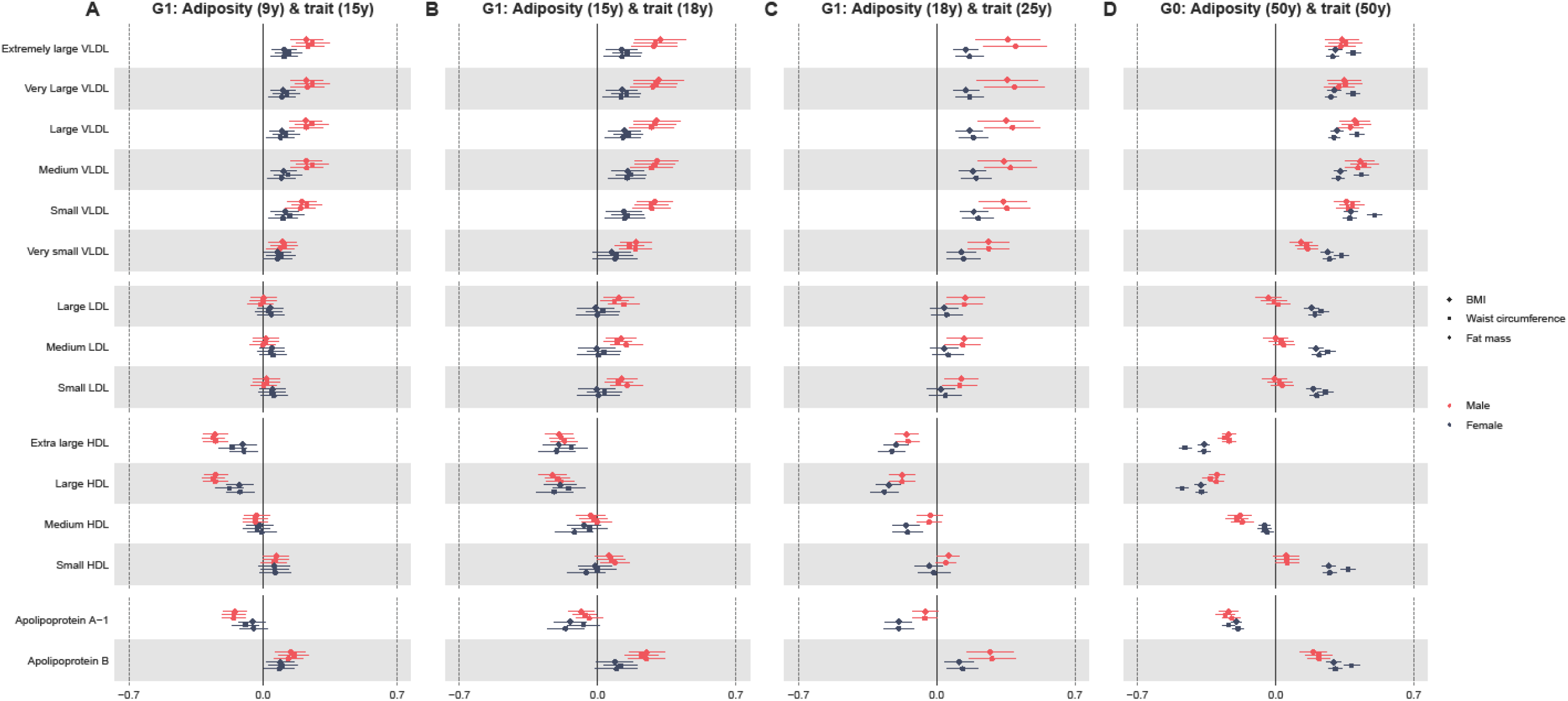
Sex-specific association of BMI, waist circumference and fat mass (per SD increase) with standardised lipoprotein concentrations from childhood to mid life. **Legend**: BMI, body mass index; G1, offspring generation 1; G0, parent generation 0; HDL, high-density lipoprotein; LDL, low-density lipoprotein; VLDL, very-low-density lipoprotein. Results shown are standardised differences in cardiometabolic trait per standard deviation increase in BMI, waist circumference and fat mass in each sex separately. G1 analyses are adjusted for age at clinic completion, ethnicity, child’s mother and father education, maternal smoking during pregnancy, birthweight, gestational age, maternal age, household social class and height and height^2^. Analyses of outcomes at 18y and 25y are also additionally adjusted for G1 offspring smoking. G0 analyses are adjusted for age at clinic completion, ethnicity, education, smoking during G1 cohort pregnancy, own social class and height and height^2^.SD of BMI = 2.7 kg/m^2^, 2.9 kg/m^2^, 3.8 kg/m^2^ and 4.8 kg/m^2^ at 9y, 15y, 18y and 50y respectively. SD of waist circumference = 7.4cm, 7.8cm and 13cm at 9y, 15y and 50y respectively. SD of fat mass = 5kg, 7.8 kg, 9.8kg and 10.2kg at 9y, 15y, 18y and 50y respectively.

Adiposity measures were positively associated with all concentrations of low-density lipoprotein (LDL) particles at 18y and 25y in males only and at 50y in females only. In contrast, adiposity measures were inversely associated with concentrations of very large and large high-density lipoprotein (HDL) particles at all ages (Figure 2) in both sexes; associations were stronger in males at 15y, similar in males and females at 18y and stronger in females at 25y and 50y. Adiposity measures were inversely associated with concentrations of medium HDL particles at 25y in females only and at 50y in both sexes; inverse associations with concentrations of medium HDL particles were stronger in males at 50y. Adiposity measures were positively associated with concentrations of small HDL particles at 50y only in both sexes; positive associations with small HDL were stronger in females.

Adiposity measures were inversely associated with apolipoprotein A-1 and positively associated with apolipoprotein B at each age in both sexes; at younger ages associations with apolipoprotein A-1 concentrations were stronger in females while associations with apolipoprotein B concentrations were stronger in males. At 50y, associations were similar between the sexes for apolipoprotein A-1 and stronger in females for apolipoprotein B.

### Associations of adiposity measures with cholesterol, triglycerides and glycaemic trait concentrations in females and males

Adiposity measures were positively associated with remnant and VLDL cholesterol in both sexes at all ages (Figure 3); associations were stronger in males at younger ages but similar between the sexes at 50y. Adiposity measures were positively associated with total, free, esterified and LDL cholesterol in males only at 18 and 25y. At 50y, adiposity measures were inversely associated with total, free, esterified and LDL cholesterol concentrations in males but positively associated with these in females.

**Figure 3.**
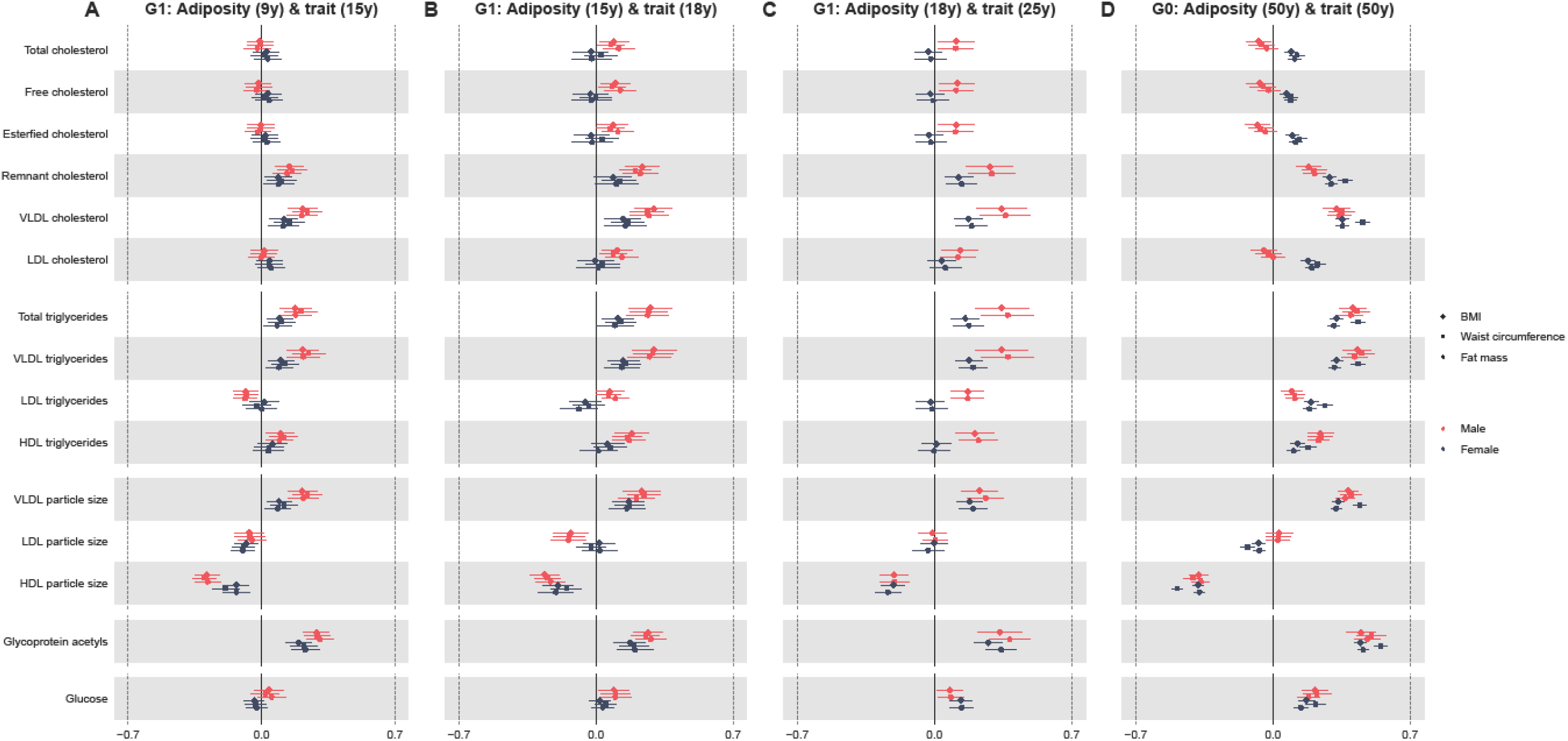
Sex-specific association of in BMI, waist circumference and fat mass (per SD increase) with standardised cholesterol and triglyceride concentrations from childhood to mid life. Legend: BMI, body mass index; G1, offspring generation 1; G0, parent generation 0; HDL, high-density lipoprotein; LDL, low-density lipoprotein; VLDL, very-low-density lipoprotein. Results shown are standardised differences in cardiometabolic trait per standard deviation increase in BMI, waist circumference and fat mass in each sex separately. G1 analyses are adjusted for age at clinic completion, ethnicity, child’s mother and father education, maternal smoking during pregnancy, birthweight, gestational age, maternal age, household social class and height and height^2^. Analyses of outcomes at 18y and 25y are also additionally adjusted for G1 offspring smoking. G0 analyses are adjusted for age at clinic completion, ethnicity, education, smoking during G1 cohort pregnancy, own social class and height and height^2^.SD of BMI = 2.7 kg/m^2^, 2.9 kg/m^2^, 3.8 kg/m^2^and 4.8 kg/m^2^at 9y, 15y, 18y and 50y respectively. SD of waist circumference = 7.4cm, 7.8cm and 13cm at 9y, 15y and 50y respectively. SD of fat mass = 5kg, 7.8 kg, 9.8kg and 10.2kg at 9y, 15y, 18y and 50y respectively.

Adiposity measures were positively associated with total and VLDL triglycerides at all ages in both sexes; associations were stronger in males at each age. For instance, a 1-SD higher BMI at 18y (SD = 3.8 kg/m^2^) was associated with 0.34 SD (95% CI = 0.21, 0.48) higher total triglycerides at 25y in males compared with 0.16 SD (95% CI = 0.09, 0.23) in females. BMI was positively associated with LDL and HDL triglycerides in males only at 18 and 25y and in both sexes at 50y.

Adiposity measures were positively associated with VLDL particle size and inversely associated with HDL particle size at all ages; associations were stronger in males compared with females at 15y and 18y while associations were similar between the sexes at 25y and 50y. Adiposity measures were positively associated with glycoprotein acetyls in both sexes at each time point and associations were stronger in males at 15y and 18y but more similar between the sexes at 25y and 50y. Adiposity measures were similarly positively associated with glucose in both sexes at all time points.

Sex-specific associations of BMI, waist circumference and fat mass with cardiometabolic traits were similar at each age (Figure 2 & 3). Unadjusted and confounder-adjusted results were also similar (eTable 4 & 5). Results in original units of traits are shown in eTable 6.

### Additional and sensitivity analyses

Our results were not appreciably different when analyses were performed using the sex-specific mean and SD of each adiposity measure for standardisation (eFigure 1 & 2) or when analyses were restricted to full family units (eFigure 3 & 4). Associations were also similar when participants in the upper fifth of the adiposity distribution were excluded from analyses suggesting that modest departures from linearity for some traits are unlikely to have impacted our results (eFigure 5 & 6).

## Discussion

In this UK prospective cohort study including two generations of participants, we examined sex-specific associations of general adiposity with detailed cardiometabolic traits measured in mid and late adolescence and early-and mid-adulthood. Overall, our findings suggest that the adverse cardiometabolic effects of adiposity are stronger and begin earlier in the life course in males compared with females until mid life. Adolescent and young adult males may therefore be high priority targets for obesity prevention efforts. Our findings also demonstrate high consistency between BMI, waist circumference, and DXA fat mass in their ability to detect associations with detailed cardiometabolic traits at each life stage, supporting the continued use of BMI as a simple and inexpensive measure of adiposity for causal analyses in both sexes across the life course.

Cardiometabolic disease risk tends to appear higher in males until mid life, after which risk becomes more similar between females and males (5). In a previous ALPSAC study of the same cardiometabolic traits studied here, absolute levels of several key causal CHD traits were higher in males compared with females in early life and became more similar or lower in males compared with females in mid life (3). Our results suggest that stronger early life and weaker later life effects of adiposity on cardiometabolic traits in males play an important role in these previously identified life course sex differences in cardiometabolic risk (3). In addition, several of the traits that were higher in males in that study (3) and more strongly associated with adiposity in males in our study such as LDL cholesterol (26), VLDL triglycerides (27) and apolipoprotein-B (28-30) are very likely causal for CHD. HDL cholesterol, an important non-causal marker of CHD risk (31) was also lower among males in that study and more strongly associated with adiposity among males in the present study. If sex differences in associations of adiposity with these cardiometabolic traits reflect sex differences in causal effects, the findings provide insights into potential drivers of sex differences in age-adjusted risk of CHD across the life course. The more adverse pattern seen among males, despite their lower total fat than females, in turn likely reflects sex differences in patterns of fat storage with males storing more metabolically harmful abdominal fat and females storing more peripheral fat which is less metabolically active. This is exemplified by the so-called ‘favourable adiposity’ phenotype, underpinned by a genotype wherein higher fat volume is accompanied by higher insulin sensitivity and better cardiometabolic health (32, 33). While previous studies have demonstrated strong comparability of observational and causal estimates of BMI and metabolites (7), additional study designs including Mendelian randomisation are required to interrogate the causality of observed sex-specific associations found here.

While studies examining associations of adiposity with cardiometabolic traits at multiple life stages are limited, our results are comparable with previous work in this (34, 35) cohort showing sex-specific associations of adiposity with conventional cardiometabolic risk factors in adolescence to the disadvantage of males. The results are also comparable to findings from a Young Finns analysis of 12,664 adolescents and young adults aged 16-39, which demonstrated similar evidence for stronger sex-specific associations of BMI with causal CHD susceptibility traits in males, using evidence from Mendelian randomisation (7); however, a key limitation was limited statistical power to estimate sex-specific effects with certainty. The similarity of sex specific associations of BMI, waist circumference and fat mass with cardiometabolic traits at each life stage reported here is comparable to previous studies in adolescence and adulthood (14, 34), though some analyses in adults have reported differences for CVD endpoints (15). However, previous work has focused predominantly on examining and comparing associations of adiposity with conventional cardiometabolic traits, often at single time points in adolescence or adulthood. Our work extends beyond this by comparing associations with 148 trait concentrations at multiple life stages in adolescence and adulthood. Importantly, the findings support the use of BMI as a general, easily measured and inexpensive method of adiposity measurement at a population level from childhood to mid adulthood, for causation purposes.

### Strengths and Limitations

There are several strengths to our study including the use of both general and central measures of adiposity and the direct comparison of the associations of these measures with ∼148 detailed cardiometabolic trait concentrations from a targeted metabolomics platform measured at multiple life stages. In addition, we have performed multiple sensitivity analyses including examining the robustness of our findings to standardisation of the exposures using sex-specific mean and SD, to potential selection issues (by performing analyses only on mother-father-offspring trios) and to threshold effects of adiposity on outcomes (by excluding the upper fifth of the adiposity distribution).

Limitations include missing data and loss to follow-up leading to modest sample sizes for inclusion in analyses compared with the numbers available in the originally recruited birth cohort. If loss to follow up is differentially related to both exposure and outcome by sex, estimates of sex-specific associations at each life stage could be biased. In addition, estimates of sex-specific associations in the G1 cohort compared with the G0 parent cohort could be further undermined by different degrees of selection bias in the G1 and G0 cohorts. For instance, although we found demographic characteristics were comparable between included and excluded females and males in each cohort separately, we did find some evidence of greater sex-specific selection in the G0 cohort, which may impact the validity of the comparison of the two cohorts. Furthermore, while measures analysed in G1 cohort analyses were prospective, analyses at 50y were cross-sectional. Importantly, we cannot exclude the possibility that differences in estimates between the two cohorts are not a product of “cohort effects” driven by generational differences between the children and their parents such as greater exposure of the G1 cohort to the current obesity epidemic. While we included and compared three general measures of adiposity here, the measures do not capture fine scale adiposity and adiposity distribution differences in females and males including shifts towards “metabolically favourable adiposity”(32) which may explain less adverse effects of adiposity among females found here. Residual confounding may also persist in some of our analyses due to lack of adjustment for other confounders such as alcohol use or medical conditions, particularly in the G0 parent cohort. Finally, participants were predominantly of White ethnicity and more socially advantaged and thus, our results may not be generalizable to other populations.

## Conclusion

The results of this study suggest that the adverse cardiometabolic effects of adiposity are stronger and begin earlier in the life course in males compared with females until mid life, particularly for key atherogenic lipids. Adolescent and young adult males may therefore be high priority targets for obesity prevention efforts.

**Table 1.**
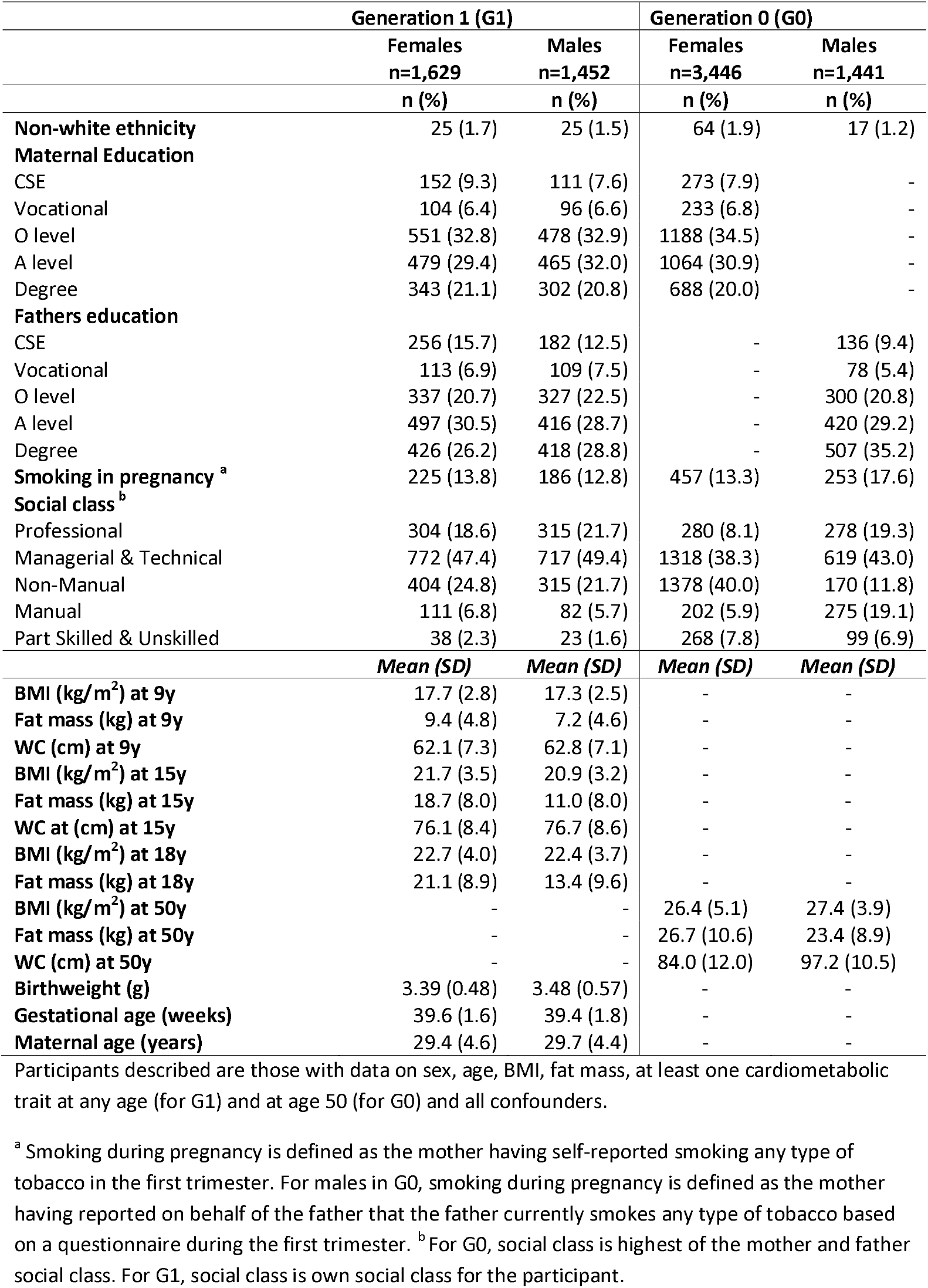
Characteristics of participants included in analyses, by sex

| **Table 1 Characteristics of participants included in analyses, by sex** |  |  |  |  |
| --- | --- | --- | --- | --- |
|  | **Generation 1 (G1)** |  | **Generation 0 (G0)** |  |
|  | **Females**<br>**n=1,629** | **Males**<br>**n=1,452** | **Females**<br>**n=3,446** | **Males**<br>**n=1,441** |
|  | **n (%)** | **n (%)** | **n (%)** | **n (%)** |
| **Non-white ethnicity** | 25 (1.7) | 25 (1.5) | 64 (1.9) | 17 (1.2) |
| **Maternal Education** |  |  |  |  |
| CSE | 152 (9.3) | 111 (7.6) | 273 (7.9) | - |
| Vocational | 104 (6.4) | 96 (6.6) | 233 (6.8) | - |
| O level | 551 (32.8) | 478 (32.9) | 1188 (34.5) | - |
| A level | 479 (29.4) | 465 (32.0) | 1064 (30.9) | - |
| Degree | 343 (21.1) | 302 (20.8) | 688 (20.0) | - |
| **Fathers education** |  |  |  |  |
| CSE | 256 (15.7) | 182 (12.5) | - | 136 (9.4) |
| Vocational | 113 (6.9) | 109 (7.5) | - | 78 (5.4) |
| O level | 337 (20.7) | 327 (22.5) | - | 300 (20.8) |
| A level | 497 (30.5) | 416 (28.7) | - | 420 (29.2) |
| Degree | 426 (26.2) | 418 (28.8) | - | 507 (35.2) |
| **Smoking in pregnancy <sup>a</sup>** | 225 (13.8) | 186 (12.8) | 457 (13.3) | 253 (17.6) |
| **Social class <sup>b</sup>** |  |  |  |  |
| Professional | 304 (18.6) | 315 (21.7) | 280 (8.1) | 278 (19.3) |
| Managerial & Technical | 772 (47.4) | 717 (49.4) | 1318 (38.3) | 619 (43.0) |
| Non-Manual | 404 (24.8) | 315 (21.7) | 1378 (40.0) | 170 (11.8) |
| Manual | 111 (6.8) | 82 (5.7) | 202 (5.9) | 275 (19.1) |
| Part Skilled & Unskilled | 38 (2.3) | 23 (1.6) | 268 (7.8) | 99 (6.9) |
|  | **Mean (SD)** | **Mean (SD)** | **Mean (SD)** | **Mean (SD)** |
| **BMI (kg/m<sup>2</sup>) at 9y** | 17.7 (2.8) | 17.3 (2.5) | - | - |
| **Fat mass (kg) at 9y** | 9.4 (4.8) | 7.2 (4.6) | - | - |
| **WC (cm) at 9y** | 62.1 (7.3) | 62.8 (7.1) | - | - |
| **BMI (kg/m<sup>2</sup>) at 15y** | 21.7 (3.5) | 20.9 (3.2) | - | - |
| **Fat mass (kg) at 15y** | 18.7 (8.0) | 11.0 (8.0) | - | - |
| **WC at (cm) at 15y** | 76.1 (8.4) | 76.7 (8.6) | - | - |
| **BMI (kg/m<sup>2</sup>) at 18y** | 22.7 (4.0) | 22.4 (3.7) | - | - |
| **Fat mass (kg) at 18y** | 21.1 (8.9) | 13.4 (9.6) | - | - |
| **BMI (kg/m<sup>2</sup>) at 50y** | - | - | 26.4 (5.1) | 27.4 (3.9) |
| **Fat mass (kg) at 50y** | - | - | 26.7 (10.6) | 23.4 (8.9) |
| **WC (cm) at 50y** | - | - | 84.0 (12.0) | 97.2 (10.5) |
| **Birthweight (g)** | 3.39 (0.48) | 3.48 (0.57) | - | - |
| **Gestational age (weeks)** | 39.6 (1.6) | 39.4 (1.8) | - | - |
| **Maternal age (years)** | 29.4 (4.6) | 29.7 (4.4) | - | - |

Participants described are those with data on sex, age, BMI, fat mass, at least one cardiometabolic trait at any age (for G1) and at age 50 (for G0) and all confounders.
<sup>a</sup> Smoking during pregnancy is defined as the mother having self-reported smoking any type of tobacco in the first trimester. For males in G0, smoking during pregnancy is defined as the mother having reported on behalf of the father that the father currently smokes any type of tobacco based on a questionnaire during the first trimester. <sup>b</sup> For G0, social class is highest of the mother and father social class. For G1, social class is own social class for the participant.

## Data Availability

Data are available upon submission and approval of a proposal to the ALSPAC Executive. Further information is available at http://www.bristol.ac.uk/alspac/researchers/our-data/.

## Acknowledgements

We are extremely grateful to all the families who took part in this study, the midwives for their help in recruiting them, and the whole ALSPAC team, which includes interviewers, computer and laboratory technicians, clerical workers, research scientists, volunteers, managers, receptionists and nurses.

