## Supplementary Material for "Sex-specific associations of adiposity with cardiometabolic traits: multi-life-stage cohort study with repeat metabolomics"

**List of Contents**

Appendix 1 Details on assessment of confounders

Table 1 Results from likelihood ratio tests of linearity testing at each time point

Table 2 Characteristics of offspring (G1 cohort) included in analyses compared to those excluded due to missing exposure, outcome or confounder data

Table 3 Characteristics of parents (G0 cohort) included in analyses compared to those excluded due to missing exposure or confounder data

Table 4 [Unadjusted sex-specific associations of BMI with concentrations of 148 cardiometabolic traits](https://uccireland-my.sharepoint.com/personal/linda_okeeffe_ucc_ie/Documents/Metabolites%20project/Write%20up/Supplementary%20Tables.xlsx) at multiple life stages

Table 5 [Adjusted sex-specific associations of BMI with 148 concentrations of cardiometabolic traits](https://uccireland-my.sharepoint.com/personal/linda_okeeffe_ucc_ie/Documents/Metabolites%20project/Write%20up/Supplementary%20Tables.xlsx) at multiple life stages

Table 6 Adjusted [[sex-specific associations of BMI with concentrations of 148 cardiometabolic traits](https://uccireland-my.sharepoint.com/personal/linda_okeeffe_ucc_ie/Documents/Metabolites%20project/Write%20up/Supplementary%20Tables.xlsx) at multiple life stages - results in original units](https://uccireland-my.sharepoint.com/personal/linda_okeeffe_ucc_ie/Documents/Metabolites%20project/Write%20up/Supplementary%20Tables.xlsx)

Figure 1: Sex-specific association of BMI, waist circumference and fat mass (per SD increase, standardised using sex-specific mean and SD) with standardised lipoprotein concentrations from childhood to mid life

Figure 2 Sex-specific association of BMI, waist circumference and fat mass (per SD increase, standardised using sex-specific mean and SD) with standardised cholesterol and triglyceride concentrations from childhood to mid life

Figure 3 Sex-specific association of BMI, waist circumference and fat mass (per SD increase) with standardised lipoprotein concentrations from childhood to mid life, excluding participants in the top fifth of the adiposity distribution

Figure 4 Sex-specific association of in BMI, waist circumference and fat mass (per SD increase) with standardised cholesterol and triglyceride concentrations from childhood to mid life, excluding participants in the top fifth of the adiposity distribution

Figure 5 Sex-specific association of BMI, waist circumference and fat mass (per SD increase) with standardised lipoprotein concentrations from childhood to mid life, including only complete family units

Figure 6 Sex-specific association of in BMI, waist circumference and fat mass (per SD increase) with standardised cholesterol and triglyceride concentrations from childhood to mid life, including only complete family units

**eAppendix 1 Details on assessment of confounders**

A questionnaire at 32 weeks gestation asked mothers to report their educational attainment, which was categorized as below O-Level (Ordinary Level; exams taken in different subjects usually at age 15-16 at the completion of legally required school attendance, equivalent to today’s UK General Certificate of Secondary Education), O-Level only, A-Level (Advanced-Level; exams taken in different subjects usually at age 18), or university degree or above. A questionnaire at 32 weeks gestation asked partners to report their educational attainment, which was categorized as below O-Level (Ordinary Level; exams taken in different subjects usually at age 15-16y at the completion of legally required school attendance, equivalent to today’s UK General Certificate of Secondary Education), O-Level only, A-Level (Advanced-Level; exams taken in different subjects usually at age 18), or university degree or above. Smoking in the first trimester of pregnancy was self-reported by mothers on their own behalf and on behalf of their partners at 18 weeks gestation. Birthweight was extracted from medical records. Gestational age at birth was estimated from clinical records. Maternal age was reported in the mother’s antenatal questionnaires. Social class was measured using data on job title and details of occupation collected about the mother and her partner from the mother’s questionnaire at 32 weeks gestation. Social class was derived using the standard occupational classification (SOC) codes developed by the United Kingdom Office of Population Census and Surveys and classified as I professional, II managerial and technical, IIINM non-manual, IIIM manual, and IV&V part skilled occupations and unskilled occupations. Household social class was measured as the highest of the mother’s or her partner’s occupational social class. Height was measured in light clothing without shoes to the nearest 0.1 cm using a Harpenden stadiometer. Smoking was measured via questionnaire at 15y and 18y in the G1 cohort.

**Table 1 Results from likelihood ratio tests of linearity testing at each time point**

|  | **BMI** | | **Waist Circumference** | | **Fat Mass** | |
| --- | --- | --- | --- | --- | --- | --- |
|  | **Females** | **Males** | **Females** | **Males** | **Females** | **Males** |
| **Adiposity measure at age 9 and metabolic trait at age 15** | | | | | | |
| Total lipids in chylomicrons and extremely large VLDL (mmol/l) | 0.114 | 0.136 | 0.197 | 0.290 | 0.173 | 0.481 |
| Total lipids in very large VLDL (mmol/l) | 0.160 | 0.117 | 0.193 | 0.166 | 0.287 | 0.480 |
| Total lipids in large VLDL (mmol/l) | 0.230 | 0.115 | 0.217 | 0.110 | 0.388 | 0.565 |
| Total lipids in medium VLDL (mmol/l) | 0.257 | 0.133 | 0.258 | 0.126 | 0.338 | 0.545 |
| Total lipids in small VLDL (mmol/l) | 0.162 | 0.345 | 0.397 | 0.421 | 0.174 | 0.566 |
| Total lipids in very small VLDL (mmol/l) | 0.553 | 0.765 | 0.955 | 0.683 | 0.235 | 0.674 |
| Total lipids in large LDL (mmol/l) | 0.405 | 0.088 | 0.546 | 0.151 | 0.461 | 0.432 |
| Total lipids in medium LDL (mmol/l) | 0.338 | 0.068 | 0.618 | 0.206 | 0.419 | 0.512 |
| Total lipids in small LDL (mmol/l) | 0.323 | 0.066 | 0.664 | 0.218 | 0.413 | 0.495 |
| Total lipids in very large HDL (mmol/l) | 0.182 | 0.023 | 0.055 | 0.145 | 0.413 | 0.003 |
| Total lipids in large HDL (mmol/l) | 0.148 | 0.029 | 0.018 | 0.077 | 0.263 | 0.004 |
| Total lipids in medium HDL (mmol/l) | 0.669 | 0.287 | 0.352 | 0.179 | 0.367 | 0.206 |
| Total lipids in small HDL (mmol/l) | 0.453 | 0.448 | 0.735 | 0.100 | 0.519 | 0.829 |
| Apolipoprotein A-I (g/l) | 0.326 | 0.017 | 0.102 | 0.131 | 0.459 | 0.006 |
| Apolipoprotein B (g/l) | 0.322 | 0.444 | 0.909 | 0.991 | 0.190 | 0.830 |
| Serum total cholesterol (mmol/l) | 0.459 | 0.104 | 0.472 | 0.168 | 0.540 | 0.215 |
| Free cholesterol (mmol/l) | 0.544 | 0.138 | 0.523 | 0.334 | 0.339 | 0.263 |
| Esterified cholesterol (mmol/l) | 0.435 | 0.101 | 0.410 | 0.108 | 0.650 | 0.208 |
| Remnant cholesterol (non-HDL, non-LDL -cholesterol) (mmol/l) | 0.565 | 0.705 | 0.983 | 0.896 | 0.218 | 0.717 |
| Total cholesterol in VLDL (mmol/l) | 0.357 | 0.327 | 0.671 | 0.627 | 0.140 | 0.557 |
| Total cholesterol in LDL (mmol/l) | 0.416 | 0.108 | 0.589 | 0.131 | 0.470 | 0.493 |
| Total cholesterol in HDL (mmol/l) | 0.271 | 0.021 | 0.066 | 0.102 | 0.311 | 0.003 |
| Serum total triglycerides (mmol/l) | 0.133 | 0.224 | 0.305 | 0.221 | 0.280 | 0.633 |
| Triglycerides in VLDL (mmol/l) | 0.188 | 0.150 | 0.194 | 0.117 | 0.354 | 0.531 |
| Triglycerides in LDL (mmol/l) | 0.208 | 0.002 | 0.326 | 0.796 | 0.457 | 0.245 |
| Triglycerides in HDL (mmol/l) | 0.496 | 0.456 | 0.811 | 0.415 | 0.465 | 0.735 |
| **Adiposity measure at age 15 and metabolic trait at age 18** | | | | | | |
| Total lipids in chylomicrons and extremely large VLDL (mmol/l) | 0.001 | 0.155 | 0.298 | 0.726 | 0.124 | 0.444 |
| Total lipids in very large VLDL (mmol/l) | 0.002 | 0.163 | 0.432 | 0.690 | 0.087 | 0.382 |
| Total lipids in large VLDL (mmol/l) | 0.003 | 0.214 | 0.500 | 0.674 | 0.063 | 0.374 |
| Total lipids in medium VLDL (mmol/l) | 0.002 | 0.407 | 0.392 | 0.715 | 0.051 | 0.467 |
| Total lipids in small VLDL (mmol/l) | 0.004 | 0.405 | 0.337 | 0.898 | 0.148 | 0.391 |
| Total lipids in very small VLDL (mmol/l) | 0.007 | 0.231 | 0.090 | 0.387 | 0.299 | 0.173 |
| Total lipids in large LDL (mmol/l) | 0.120 | 0.450 | 0.360 | 0.291 | 0.332 | 0.187 |
| Total lipids in medium LDL (mmol/l) | 0.107 | 0.503 | 0.378 | 0.394 | 0.328 | 0.204 |
| Total lipids in small LDL (mmol/l) | 0.083 | 0.565 | 0.361 | 0.435 | 0.325 | 0.192 |
| Total lipids in very large HDL (mmol/l) | 0.347 | 0.804 | 0.030 | 0.543 | 0.517 | 0.287 |
| Total lipids in large HDL (mmol/l) | 0.040 | 0.973 | 0.002 | 0.384 | 0.426 | 0.601 |
| Total lipids in medium HDL (mmol/l) | 0.040 | 0.969 | 0.014 | 0.400 | 0.865 | 0.851 |
| Total lipids in small HDL (mmol/l) | 0.019 | 0.689 | 0.104 | 0.514 | 0.989 | 0.451 |
| Apolipoprotein A-I (g/l) | 0.064 | 0.755 | 0.015 | 0.225 | 0.505 | 0.817 |
| Apolipoprotein B (g/l) | 0.009 | 0.444 | 0.266 | 0.737 | 0.114 | 0.449 |
| Serum total cholesterol (mmol/l) | 0.084 | 0.541 | 0.418 | 0.209 | 0.300 | 0.199 |
| Free cholesterol (mmol/l) | 0.153 | 0.419 | 0.565 | 0.254 | 0.191 | 0.120 |
| Esterified cholesterol (mmol/l) | 0.071 | 0.624 | 0.394 | 0.157 | 0.408 | 0.248 |
| Remnant cholesterol (non-HDL, non-LDL -cholesterol) (mmol/l) | 0.006 | 0.414 | 0.162 | 0.542 | 0.149 | 0.360 |
| Total cholesterol in VLDL (mmol/l) | 0.001 | 0.498 | 0.132 | 0.839 | 0.110 | 0.559 |
| Total cholesterol in LDL (mmol/l) | 0.150 | 0.509 | 0.341 | 0.297 | 0.304 | 0.172 |
| Total cholesterol in HDL (mmol/l) | 0.029 | 0.872 | 0.003 | 0.319 | 0.421 | 0.807 |
| Serum total triglycerides (mmol/l) | 0.009 | 0.135 | 0.836 | 0.735 | 0.109 | 0.404 |
| Triglycerides in VLDL (mmol/l) | 0.005 | 0.214 | 0.518 | 0.674 | 0.074 | 0.351 |
| Triglycerides in LDL (mmol/l) | 0.019 | 0.033 | 0.200 | 0.895 | 0.759 | 0.541 |
| Triglycerides in HDL (mmol/l) | 0.044 | 0.269 | 0.619 | 0.797 | 0.267 | 0.913 |
| **Adiposity measure at age 18 and metabolic trait at age 25** | | | | | | |
| Total lipids in chylomicrons and extremely large VLDL (mmol/l) | 0.170 | 0.567 | 0.158 | 0.251 | - | - |
| Total lipids in very large VLDL (mmol/l) | 0.134 | 0.614 | 0.123 | 0.317 | - | - |
| Total lipids in large VLDL (mmol/l) | 0.082 | 0.615 | 0.132 | 0.293 | - | - |
| Total lipids in medium VLDL (mmol/l) | 0.062 | 0.528 | 0.131 | 0.315 | - | - |
| Total lipids in small VLDL (mmol/l) | 0.105 | 0.302 | 0.313 | 0.321 | - | - |
| Total lipids in very small VLDL (mmol/l) | 0.555 | 0.297 | 0.999 | 0.584 | - | - |
| Total lipids in large LDL (mmol/l) | 0.722 | 0.805 | 0.772 | 0.919 | - | - |
| Total lipids in medium LDL (mmol/l) | 0.871 | 0.687 | 0.724 | 0.868 | - | - |
| Total lipids in small LDL (mmol/l) | 0.921 | 0.642 | 0.570 | 0.823 | - | - |
| Total lipids in very large HDL (mmol/l) | 0.227 | 0.057 | 0.002 | 0.382 | - | - |
| Total lipids in large HDL (mmol/l) | 0.145 | 0.135 | 0.010 | 0.308 | - | - |
| Total lipids in medium HDL (mmol/l) | 0.241 | 0.392 | 0.231 | 0.104 | - | - |
| Total lipids in small HDL (mmol/l) | 0.593 | 0.088 | 0.070 | 0.192 | - | - |
| Apolipoprotein A-I (g/l) | 0.269 | 0.624 | 0.110 | 0.250 | - | - |
| Apolipoprotein B (g/l) | 0.738 | 0.543 | 0.980 | 0.704 | - | - |
| Serum total cholesterol (mmol/l) | 0.757 | 0.916 | 0.465 | 0.778 | - | - |
| Free cholesterol (mmol/l) | 0.940 | 0.856 | 0.666 | 0.850 | - | - |
| Esterified cholesterol (mmol/l) | 0.751 | 0.936 | 0.469 | 0.747 | - | - |
| Remnant cholesterol (non-HDL, non-LDL -cholesterol) (mmol/l) | 0.663 | 0.466 | 0.981 | 0.641 | - | - |
| Total cholesterol in VLDL (mmol/l) | 0.276 | 0.320 | 0.644 | 0.308 | - | - |
| Total cholesterol in LDL (mmol/l) | 0.787 | 0.764 | 0.761 | 0.897 | - | - |
| Total cholesterol in HDL (mmol/l) | 0.119 | 0.318 | 0.051 | 0.151 | - | - |
| Serum total triglycerides (mmol/l) | 0.124 | 0.538 | 0.274 | 0.333 | - | - |
| Triglycerides in VLDL (mmol/l) | 0.058 | 0.556 | 0.115 | 0.320 | - | - |
| Triglycerides in LDL (mmol/l) | 0.930 | 0.600 | 0.729 | 0.514 | - | - |
| Triglycerides in HDL (mmol/l) | 0.856 | 0.691 | 0.192 | 0.249 | - | - |
| **Adiposity measure at age 50 and metabolic trait at age 50** | | | | | | |
| Total lipids in chylomicrons and extremely large VLDL (mmol/l) | 0.00004 | 0.417 | 0.0003 | 0.361 | 0.177 | 0.799 |
| Total lipids in very large VLDL (mmol/l) | 0.00002 | 0.577 | 0.0002 | 0.308 | 0.111 | 0.808 |
| Total lipids in large VLDL (mmol/l) | 0.00002 | 0.562 | 0.0003 | 0.058 | 0.282 | 0.528 |
| Total lipids in medium VLDL (mmol/l) | 0.00008 | 0.681 | 0.004 | 0.009 | 0.740 | 0.592 |
| Total lipids in small VLDL (mmol/l) | 0.007 | 0.413 | 0.098 | 0.0002 | 0.554 | 0.659 |
| Total lipids in very small VLDL (mmol/l) | 0.616 | 0.026 | 0.787 | 0.0001 | 0.006 | 0.005 |
| Total lipids in large LDL (mmol/l) | 0.960 | 0.006 | 0.472 | 0.003 | 0.040 | 0.001 |
| Total lipids in medium LDL (mmol/l) | 0.987 | 0.016 | 0.414 | 0.006 | 0.062 | 0.002 |
| Total lipids in small LDL (mmol/l) | 0.984 | 0.020 | 0.379 | 0.007 | 0.049 | 0.002 |
| Total lipids in very large HDL (mmol/l) | 0.387 | 0.348 | 0.046 | 0.0002 | 0.123 | 0.917 |
| Total lipids in large HDL (mmol/l) | 0.056 | 0.497 | 0.064 | 0.001 | 0.166 | 0.654 |
| Total lipids in medium HDL (mmol/l) | 0.294 | 0.489 | 0.382 | 0.910 | 0.406 | 0.209 |
| Total lipids in small HDL (mmol/l) | 0.799 | 0.311 | 0.042 | 0.349 | 0.575 | 0.325 |
| Apolipoprotein A-I (g/l) | 0.369 | 0.377 | 0.015 | 0.887 | 0.386 | 0.076 |
| Apolipoprotein B (g/l) | 0.398 | 0.124 | 0.254 | 0.0001 | 0.143 | 0.073 |
| Serum total cholesterol (mmol/l) | 0.976 | 0.017 | 0.263 | 0.017 | 0.080 | 0.001 |
| Free cholesterol (mmol/l) | 0.988 | 0.041 | 0.249 | 0.031 | 0.246 | 0.003 |
| Esterified cholesterol (mmol/l) | 0.948 | 0.013 | 0.140 | 0.020 | 0.041 | 0.001 |
| Remnant cholesterol (non-HDL, non-LDL -cholesterol) (mmol/l) | 0.292 | 0.137 | 0.380 | 0.0002 | 0.090 | 0.049 |
| Total cholesterol in VLDL (mmol/l) | 0.009 | 0.470 | 0.121 | 0.0003 | 0.394 | 0.338 |
| Total cholesterol in LDL (mmol/l) | 0.957 | 0.005 | 0.442 | 0.003 | 0.027 | 0.001 |
| Total cholesterol in HDL (mmol/l) | 0.046 | 0.500 | 0.085 | 0.131 | 0.282 | 0.153 |
| Serum total triglycerides (mmol/l) | 0.000 | 0.710 | 0.002 | 0.010 | 0.530 | 0.614 |
| Triglycerides in VLDL (mmol/l) | 0.000 | 0.652 | 0.002 | 0.015 | 0.539 | 0.529 |
| Triglycerides in LDL (mmol/l) | 0.772 | 0.151 | 0.128 | 0.009 | 0.682 | 0.068 |
| Triglycerides in HDL (mmol/l) | 0.024 | 0.742 | 0.001 | 0.060 | 0.185 | 0.502 |

BMI, body mass index **;** HDL, high-density lipoprotein cholesterol; LDL, low-density lipoprotein cholesterol; VLDL, very-low-density lipoprotein cholesterol.

**Table 2 Characteristics of offspring (G1 cohort) included in analyses compared to those excluded due to missing exposure, outcome or confounder data**

|  | **Female participants included**  **n= 1,629** | **Female participants excluded**  **n=4,108-5,077 ^a^** | **Male participants included**  **n= 1,452** | **Male participants excluded**  **n=4,705-5,856^a^** |
| --- | --- | --- | --- | --- |
|  | **n (%)** | **n (%)** | **n (%)** | **n (%)** |
| **Non-white ethnicity** | 25 (1.5) | 127 (2.9) | 25 (1.7) | 147 (3.0) |
| **Maternal Education** |  |  |  |  |
| CSE | 152 (9.3) | 1047 (23.8) | 111 (7.6) | 1210 (24.3) |
| Vocational | 104 (6.4) | 483 (11.0) | 96 (6.6) | 545 (10.9) |
| O level | 551 (32.8) | 1535 (34.9) | 478 (32.9) | 1753 (35.2) |
| A level | 479 (29.4) | 882 (20.1) | 465 (32.0) | 968 (19.4) |
| Degree | 343 (21.1) | 451 (10.3) | 302 (20.8) | 510 (10.2) |
| **Mothers partners education** |  |  |  |  |
| CSE | 256 (15.7) | 1263 (30.2) | 182 (12.5) | 1428 (30.3) |
| Vocational | 113 (6.9) | 389 (9.3) | 109 (7.5) | 402 (8.5) |
| O level | 337 (20.7) | 881 (21.1) | 327 (22.5) | 1006 (21.3) |
| A level | 497 (30.5) | 1044 (25.0) | 416 (28.7) | 1159 (24.6) |
| Degree | 426 (26.2) | 608 (14.5) | 418 (28.8) | 723 (15.3) |
| **Maternal pregnancy smoking** | 225 (13.8) | 1326 (27.7) | 186 (12.8) | 1597 (29.6) |
| **Household social class** |  |  |  |  |
| Professional | 304 (18.6) | 426 (10.7) | 315 (21.7) | 492 (11.0) |
| Managerial & Technical | 772 (47.4) | 1584 (39.8) | 717 (49.4) | 1749 (39.0) |
| Non-Manual | 404 (24.8) | 1026 (25.8) | 315 (21.7) | 1200 (26.7) |
| Manual | 111 (6.8) | 643 (16.1) | 82 (5.7) | 725 (16.2) |
| Part Skilled & Unskilled | 38 (2.3) | 306 (7.7) | 23 (1.6) | 324 (7.2) |
|  | **Mean (SD)** | ***Mean (SD)*** | ***Mean (SD)*** | ***Mean (SD)*** |
| ***Age 9 ^b^*** |  |  |  |  |
| **BMI (kg/m^2^)** | 17.7 (2.8) | 18.1 (3.2) | 17.3 (2.5) | 17.6 (2.9) |
| **Fat mass (kg)** | 9.4 (4.8) | 10.0 (5.3) | 7.2 (4.6) | 7.5 (5.0) |
| **WC (cm)** | 62.1 (7.3) | 63.0 (8.4) | 62.8 (7.1) | 63.4 (8.0) |
| ***Age 15 ^b^*** |  |  |  |  |
| **BMI (kg/m^2^)** | 21.7 (3.5) | 22.0 (3.9) | 20.9 (3.1) | 21.2 (3.6) |
| **Fat mass (kg)** | 18.7 (8.0) | 19.3 (8.8) | 11.0 (8.0) | 11.8 (8.9) |
| **WC at (cm)** | 76.7 (8.6) | 76.9 (9.5) | 76.1 (8.4) | 77.2 (9.5) |
| ***Age 18 ^b^*** |  |  |  |  |
| **BMI (kg/m^2^)** | 22.7 (4.0) | 23.4 (4.9) | 22.4 (3.7) | 23.0 (4.1) |
| **Fat mass (kg)** | 21.1 (8.9) | 22.4 (10.4 | 13.4 (9.6) | 14.8 (10.4)) |
| **Birthweight (g)** | 3.39 (0.48) | 3.31 (0.56) | 3.48 (0.57) | 3.42 (0.62) |
| **Gestational age (weeks)** | 39.6 (1.6) | 39.3 (2.6) | 39.4 (1.8) | 39.1 (2.7) |
| **Maternal age (years)** | 29.5 (4.3) | 27.8 (4.8) | 29.7 (4.3) | 28.0 (5.0) |

^a^ Denominators for excluded participants in this table vary due to missing data for characteristics shown.

**^b^ Sample sizes vary due to missing data at each time point.**

**Table 3 Characteristics of parents (G0 cohort) included in analyses compared to those excluded due to missing exposure or confounder data**

|  | **Female participants included**  **n=3,446** | **Female participants excluded**  **n=6,540-9,730 ^a^** | **Male participants included**  **n=1,441** | **Male participants excluded**  **n=9,449-10,767 ^a^** |
| --- | --- | --- | --- | --- |
|  | **n (%)** | **n (%)** | **n (%)** | **n (%)** |
| **Non-white ethnicity** | 64 (1.9) | 257 (2.9) | 17 (1.2) | 459 (4.4) |
| **Education** |  |  |  |  |
| CSE | 273 (7.9) | 2220 (25.0) | 136 (9.4) | 2961 (28.5) |
| Vocational | 233 (6.8) | 982 (11.1) | 78 (5.4) | 923 (8.9) |
| O level | 1188 (34.5) | 3079 (34.7) | 300 (20.8) | 2216 (21.3) |
| A level | 1064 (30.9) | 1700 (19.2) | 420 (29.2) | 2662 (25.6) |
| Degree | 688 (20.0) | 898 (10.1) | 507 (35.2) | 1642 (15.8) |
| **Smoking in pregnancy ^b^** | 457 (13.3) | 2856 (29.4) | 253 (17.6) | 4113 (38.2) |
| **Household social class** |  |  |  |  |
| Professional | 280 (8.1) | 310 (4.7) | 278 (19.3) | 915 (9.7) |
| Managerial & Technical | 1318 (38.3) | 1820 (27.8) | 619 (43.0) | 3076 (32.6) |
| Non-Manual | 1378 (40.0) | 2891 (44.2) | 170 (11.8) | 1015 (10.7) |
| Manual | 202 (5.9) | 580 (8.9) | 275 (19.1) | 3142 (33.3) |
| Part Skilled & Unskilled | 268 (7.8) | 939 (14.4) | 99 (6.9) | 1301 (13.8) |
|  | ***Mean (SD)*** | ***Mean (SD)*** | ***Mean (SD)*** | ***Mean (SD)*** |
| **BMI (kg/m^2^)** | 26.4 (5.1) | 28.2 (4.5) | 27.4 (3.9) | 27.3 (6.0) |
| **Fat mass (kg)** | 26.7 (10.6) | 24.8 (9.8) | 23.4 (8.9) | 28.4 (11.6) |
| **WC (cm)** | 84.0 (12.0) | 99.0 (11.9) | 97.2 (10.5) | 86.4 (13.6) |

^a^ Denominators for excluded participants in this table vary due to missing data for characteristics shown.

^b^ Smoking during pregnancy is defined as the mother having self-reported smoking any type of tobacco in the first trimester. For males in G0, smoking during pregnancy is defined as the mother having reported on behalf of the father that the father currently smokes any type of tobacco during the first trimester.

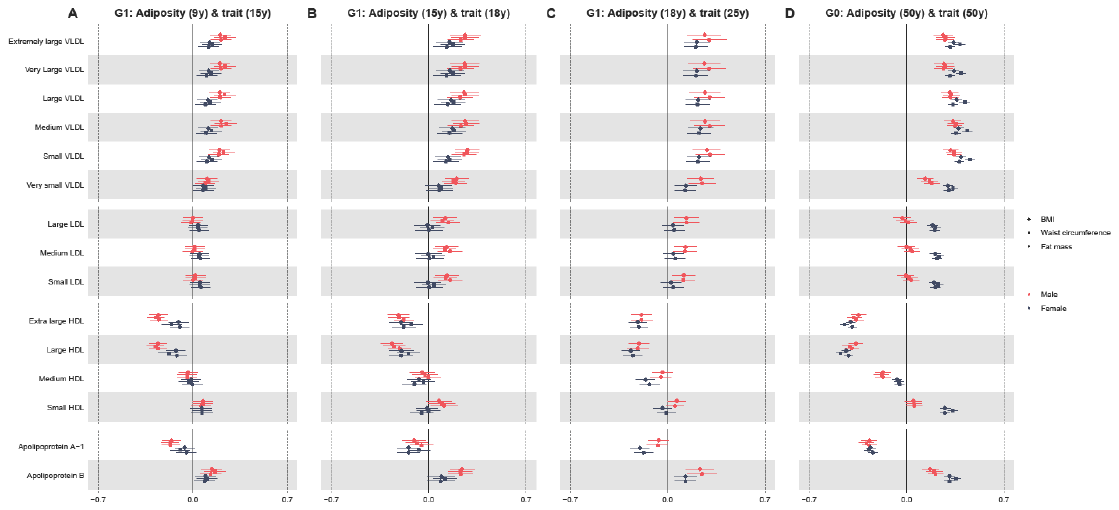
**Figure 1: Sex-specific association of BMI, waist circumference and fat mass (per SD increase, standardised using sex-specific mean and SD) with standardised lipoprotein concentrations from childhood to mid life. Legend:** BMI, body mass index; G1, offspring generation 1; G0, parent generation 0; HDL, high-density lipoprotein; LDL, low-density lipoprotein; VLDL, very-low-density lipoprotein. Results shown are standardised differences in cardiometabolic trait per standard deviation increase in BMI, waist circumference and fat mass in each sex separately. G1 analyses are adjusted for age at clinic completion, ethnicity, child’s mother and father education, [maternal](https://www.sciencedirect.com/topics/medicine-and-dentistry/gravidity-and-parity) smoking during pregnancy, birthweight, gestational age, maternal age, household social class and height and height^2^. Analyses of outcomes at 18y and 25y are also additionally adjusted for G1 offspring smoking. G0 analyses are adjusted for age at clinic completion, ethnicity, education, smoking during G1 cohort pregnancy, own social class and height and height^2^.

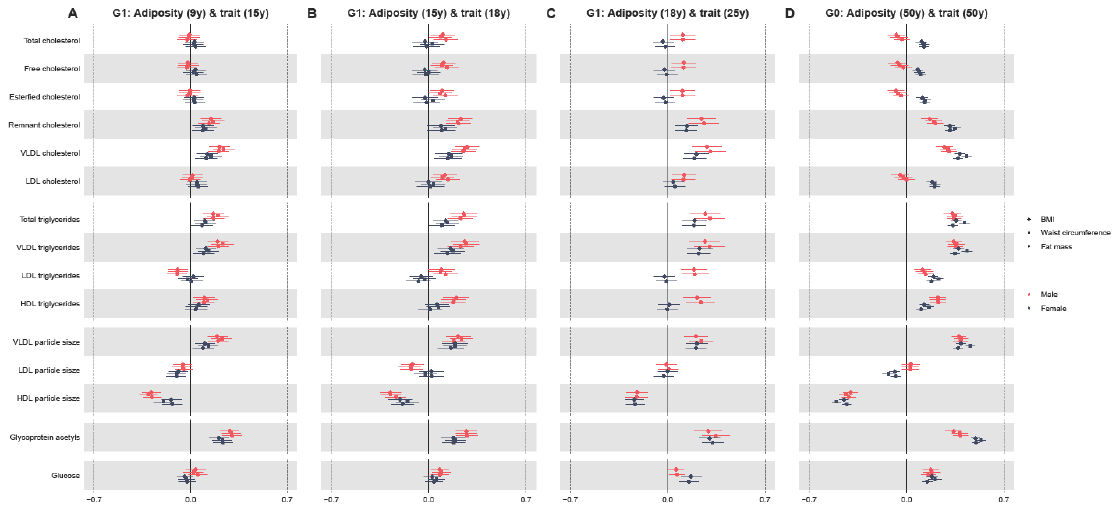
**Figure 2** **Sex-specific association of BMI, waist circumference and fat mass (per SD increase, standardised using sex-specific mean and SD) with standardised cholesterol and triglyceride concentrations from childhood to mid life. Legend:** BMI, body mass index; G1, offspring generation 1; G0, parent generation 0; HDL, high-density lipoprotein; LDL, low-density lipoprotein; VLDL, very-low-density lipoprotein. Results shown are standardised differences in cardiometabolic trait per standard deviation increase in BMI, waist circumference and fat mass in each sex separately. G1 analyses are adjusted for age at clinic completion, ethnicity, child’s mother and father education, [maternal](https://www.sciencedirect.com/topics/medicine-and-dentistry/gravidity-and-parity) smoking during pregnancy, birthweight, gestational age, maternal age, household social class and height and height^2^. Analyses of outcomes at 18y and 25y are also additionally adjusted for G1 offspring smoking. G0 analyses are adjusted for age at clinic completion, ethnicity, education, smoking during G1 cohort pregnancy, own social class and height and height^2^.

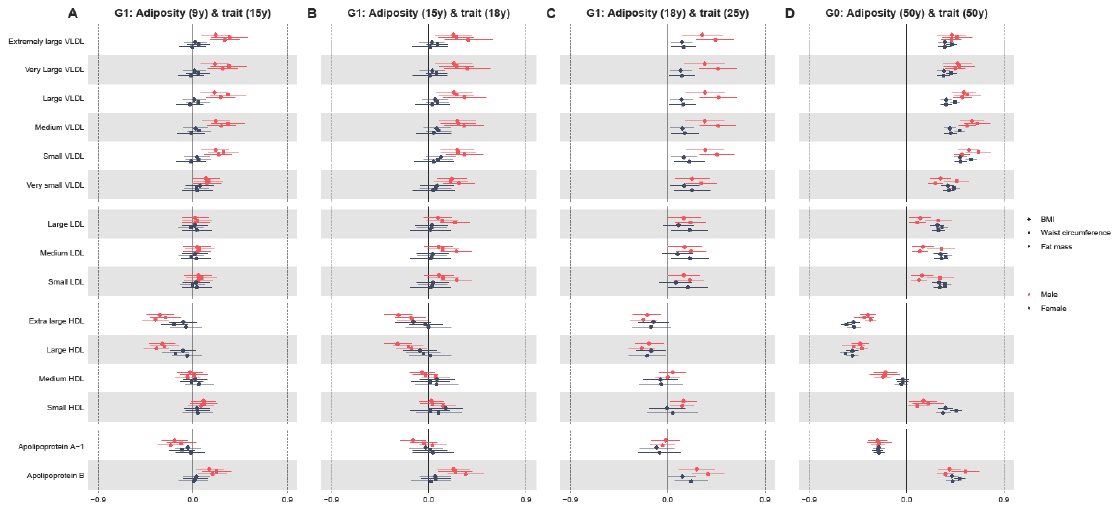
**Figure 3 Sex-specific association of BMI, waist circumference and fat mass (per SD increase) with standardised lipoprotein concentrations from childhood to mid life, excluding participants in the top fifth of the adiposity distribution. Legend:** BMI, body mass index; G1, offspring generation 1; G0, parent generation 0; HDL, high-density lipoprotein; LDL, low-density lipoprotein; VLDL, very-low-density lipoprotein. Results shown are standardised differences in cardiometabolic trait per standard deviation increase in BMI, waist circumference and fat mass in each sex separately. G1 analyses are adjusted for age at clinic completion, ethnicity, child’s mother and father education, [maternal](https://www.sciencedirect.com/topics/medicine-and-dentistry/gravidity-and-parity) smoking during pregnancy, birthweight, gestational age, maternal age, household social class and height and height^2^. Analyses of outcomes at 18y and 25y are also additionally adjusted for G1 offspring smoking. G0 analyses are adjusted for age at clinic completion, ethnicity, education, smoking during G1 cohort pregnancy, own social class and height and height^2^.

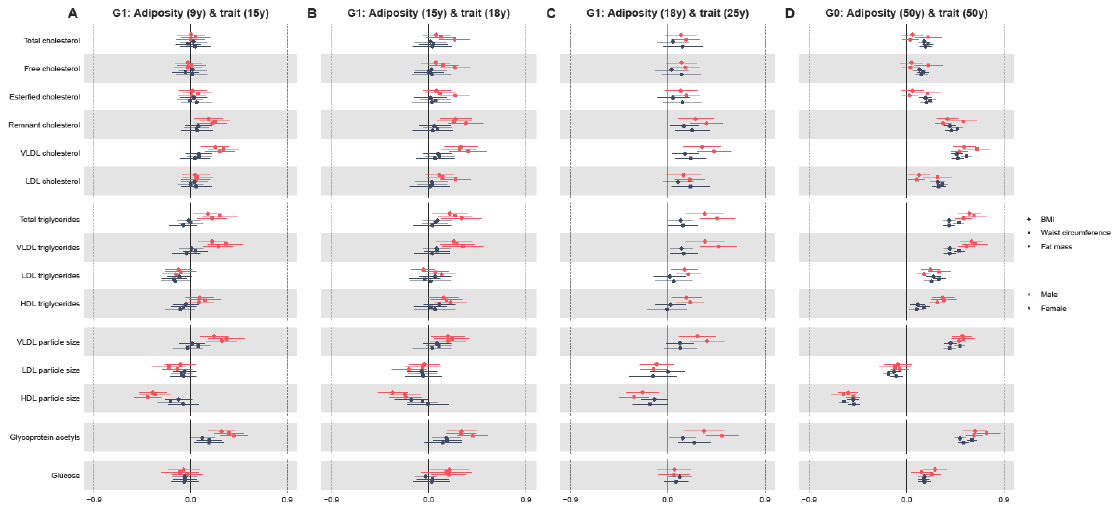
**Figure 4 Sex-specific association of in BMI, waist circumference and fat mass (per SD increase) with standardised cholesterol and triglyceride concentrations from childhood to mid life, excluding participants in the top fifth of the adiposity distribution. Legend:** BMI, body mass index; G1, offspring generation 1; G0, parent generation 0; HDL, high-density lipoprotein; LDL, low-density lipoprotein; VLDL, very-low-density lipoprotein. Results shown are standardised differences in cardiometabolic trait per standard deviation increase in BMI, waist circumference and fat mass in each sex separately. G1 analyses are adjusted for age at clinic completion, ethnicity, child’s mother and father education, [maternal](https://www.sciencedirect.com/topics/medicine-and-dentistry/gravidity-and-parity) smoking during pregnancy, birthweight, gestational age, maternal age, household social class and height and height^2^. Analyses of outcomes at 18y and 25y are also additionally adjusted for G1 offspring smoking. G0 analyses are adjusted for age at clinic completion, ethnicity, education, smoking during G1 cohort pregnancy, own social class and height and height^2^.

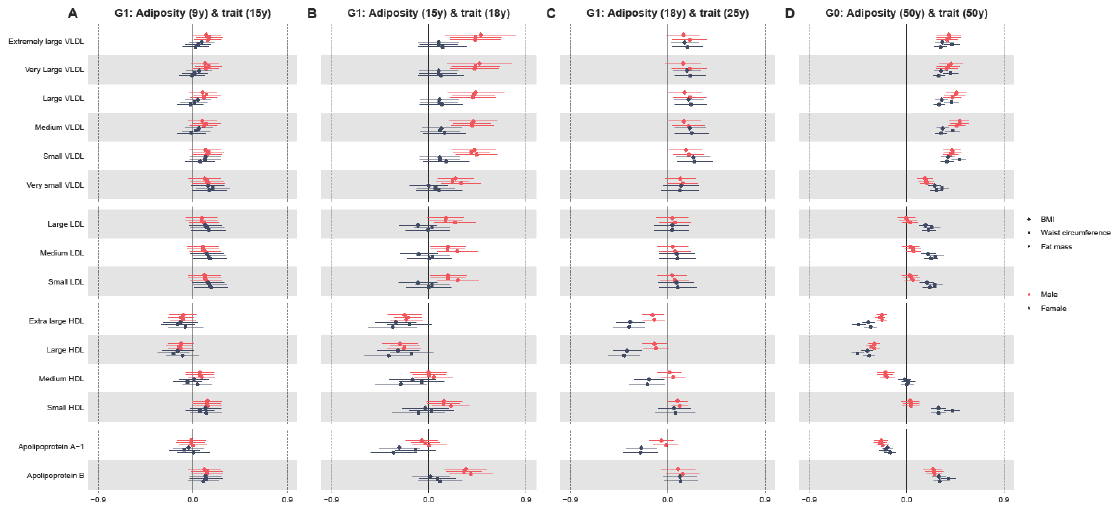
**Figure 5 Sex-specific association of BMI, waist circumference and fat mass (per SD increase) with standardised lipoprotein concentrations from childhood to mid life, including only complete family units. Legend:** BMI, body mass index; G1, offspring generation 1; G0, parent generation 0; HDL, high-density lipoprotein; LDL, low-density lipoprotein; VLDL, very-low-density lipoprotein. Results shown are standardised differences in cardiometabolic trait per standard deviation increase in BMI, waist circumference and fat mass in each sex separately. G1 analyses are adjusted for age at clinic completion, ethnicity, child’s mother and father education, [maternal](https://www.sciencedirect.com/topics/medicine-and-dentistry/gravidity-and-parity) smoking during pregnancy, birthweight, gestational age, maternal age, household social class and height and height^2^. Analyses of outcomes at 18y and 25y are also additionally adjusted for G1 offspring smoking. G0 analyses are adjusted for age at clinic completion, ethnicity, education, smoking during G1 cohort pregnancy, own social class and height and height^2^.

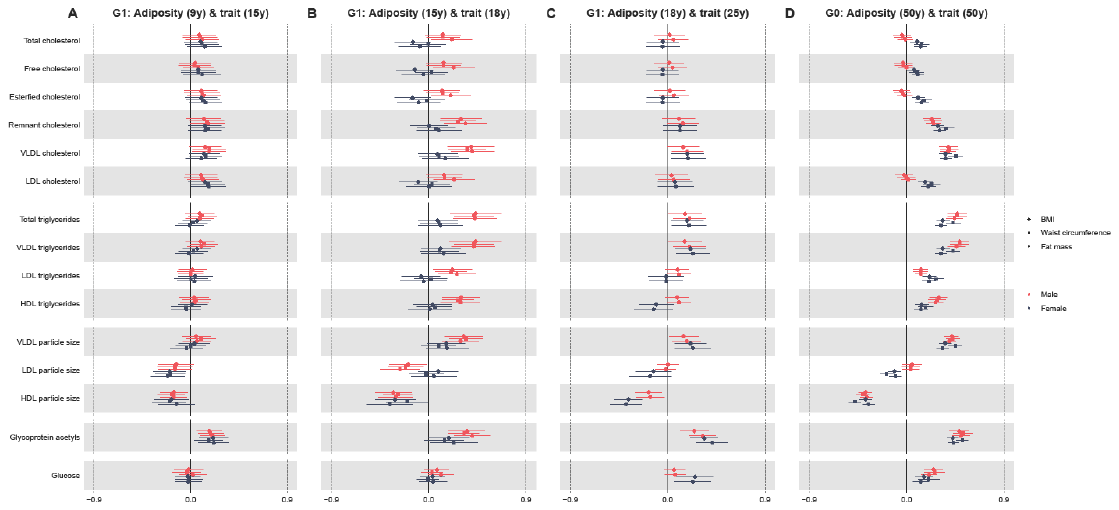
**Figure 6 Sex-specific association of in BMI, waist circumference and fat mass (per SD increase) with standardised cholesterol and triglyceride concentrations from childhood to mid life, including only complete family units. Legend:** BMI, body mass index; G1, offspring generation 1; G0, parent generation 0; HDL, high-density lipoprotein; LDL, low-density lipoprotein; VLDL, very-low-density lipoprotein. Results shown are standardised differences in cardiometabolic trait per standard deviation increase in BMI, waist circumference and fat mass in each sex separately. G1 analyses are adjusted for age at clinic completion, ethnicity, child’s mother and father education, [maternal](https://www.sciencedirect.com/topics/medicine-and-dentistry/gravidity-and-parity) smoking during pregnancy, birthweight, gestational age, maternal age, household social class and height and height^2^. Analyses of outcomes at 18y and 25y are also additionally adjusted for G1 offspring smoking. G0 analyses are adjusted for age at clinic completion, ethnicity, education, smoking during G1 cohort pregnancy, own social class and height and height^2^.
